# The unexpected dynamics of COVID-19 in Manaus, Brazil: Was herd immunity achieved?

**DOI:** 10.1101/2021.02.18.21251809

**Authors:** Daihai He, Yael Artzy-Randrup, Salihu S. Musa, Tiago Gräf, Felipe Naveca, Lewi Stone

**Affiliations:** Department of Applied Mathematics, Hong Kong Polytechnic University, Hong Kong, China; Department of Theoretical and Computational Ecology, IBED, University of Amsterdam, Amsterdam, Netherlands; Instituto Gonçalo Moniz, Fiocruz, Salvador, Bahia, Brazil; Instituto Leônidas e Maria Deane, Fiocruz, Manaus, Brazil; Mathematical Sciences, School of Science, RMIT University, Melbourne, Australia; Biomathematics Unit, School of Zoology, Faculty of Life Sciences, Tel Aviv University, Tel Aviv, Israel

## Abstract

In late March 2020, SARS-CoV-2 arrived in Manaus, Brazil, and rapidly developed into a large-scale epidemic that collapsed the local health system, and resulted in extreme death rates. Several key studies reported that ∼76% of residents of Manaus were infected (attack rate AR≃76%) by October 2020, suggesting protective herd immunity had been reached. Despite this, in November an unexpected second wave of COVID-19 struck again, and proved to be larger than the first creating a catastrophe for the unprepared population. It has been suggested that this could only be possible if the second wave was driven by reinfections. Here we use novel methods to model the epidemic from mortality data, evaluate the impact of interventions, in order to provide an alternative explanation as to why the second wave appeared. The method fits a “flexible” reproductive number *R*_0_(*t*) that changes over the epidemic, and found AR≃30-34% by October 2020, for the first wave, which is far less than required for herd immunity, yet in-line with recent seroprevalence estimates. The two-strain model provides an accurate fit to observed epidemic datasets, and finds AR≃70% by March 2021. Using genomic data, the model estimates transmissibility of the new P.1 virus lineage, as 1.9 times as transmissible as the non-P1. The model thus provides a reasonable explanation for the two-wave dynamics in Manaus, without the need to rely on reinfections which until now have only been found in small numbers in recent surveillance efforts.

**Significance:** This paper explores the concept of herd immunity and approaches for assessing attack rate during the explosive outbreak of COVID-19 in the city of Manaus, Brazil. The event has been repeatedly used to exemplify the epidemiological dynamics of the disease and the phenomenon of herd immunity, as claimed to be achieved by the end of the first wave in October 2020. A novel modelling approach reconstructs these events, specifically in the presence of interventions. The analysis finds herd immunity was far from being attained, and thus a second wave was readily possible, as tragically occurred in reality. Based on genomic data, the multi-strain model gives insights on the new highly transmissible variant of concern P.1 and role of reinfection.

## Introduction

The arrival of SARS-CoV-2 in Manaus, Brazil, in late March 2020, in short time led to an explosively growing epidemic resulting in death rates so extreme, that it captured worldwide attention and concern. The situation was characterised by the failure and breakdown of the local health system, and the appearance of mass grave sites as the number of COVID-19 deaths became unmanageable (1). Moreover, local Amazonian indigenous communities often went completely without medical services, to the point where international warnings of genocide were issued. Soon after the peak of the epidemic, in mid-May 2020, the EPICOVID19 survey estimated that 12.7% of the Manaus population had antibodies against SARS-CoV-2 (2). The epidemic proceeded to rapidly decline and was then followed by a lull from July (see Fig. 1). By October 2020, a key study (3) estimated that, approximately 76% of the city’s population had already become infected. An “Attack Rate” of this magnitude (AR∼76%) indicated that herd immunity had been achieved. As such, the population should have been protected and safe from further major COVID-19 outbreaks, at least for a reasonably lengthy period into the future. However, in reality this just provided a false sense of security. Instead, in December 2020, a vicious unexpected second wave of COVID-19 erupted resulting in the deaths of more than 170 residents per day, and rapidly triggering yet another collapse in the city’s rundown healthcare system. Alarmingly the second COVID-19 wave proved to be even larger than the first in terms of the number of COVID related deaths and number of people clinically infected (1, 4). Moreover, the same two-wave epidemic dynamics was duplicated over the whole state of Amazonas, of which Manaus is the capital and largest city (see daily death numbers in Fig. 1), and was similar in many other cities of Brazil as well, as shown in Fig. 2.

**Figure 1.**
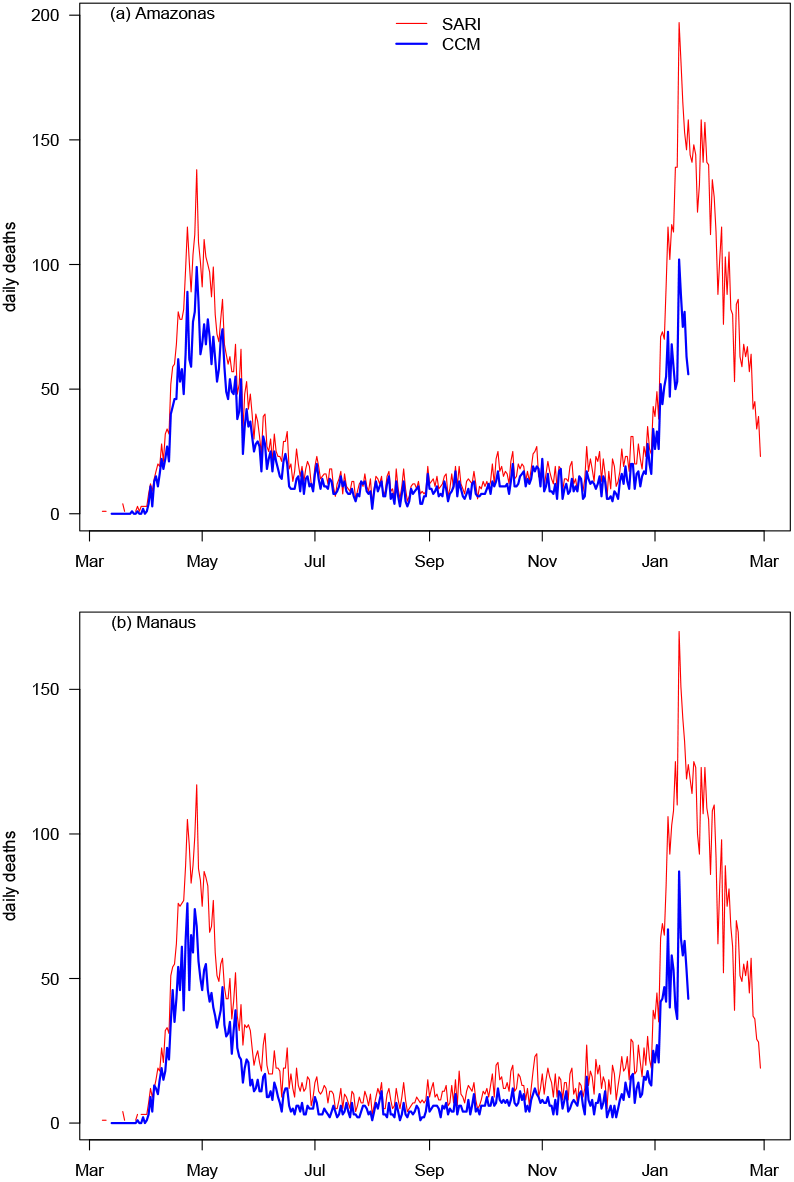
Comparison of confirmed Covid-19 mortality data (CCM) and SARI mortality datasets. Daily deaths in (a) state of Amazonas and (b) Manaus, capital city of Amazonas, from March 2020.

**Figure 2.**
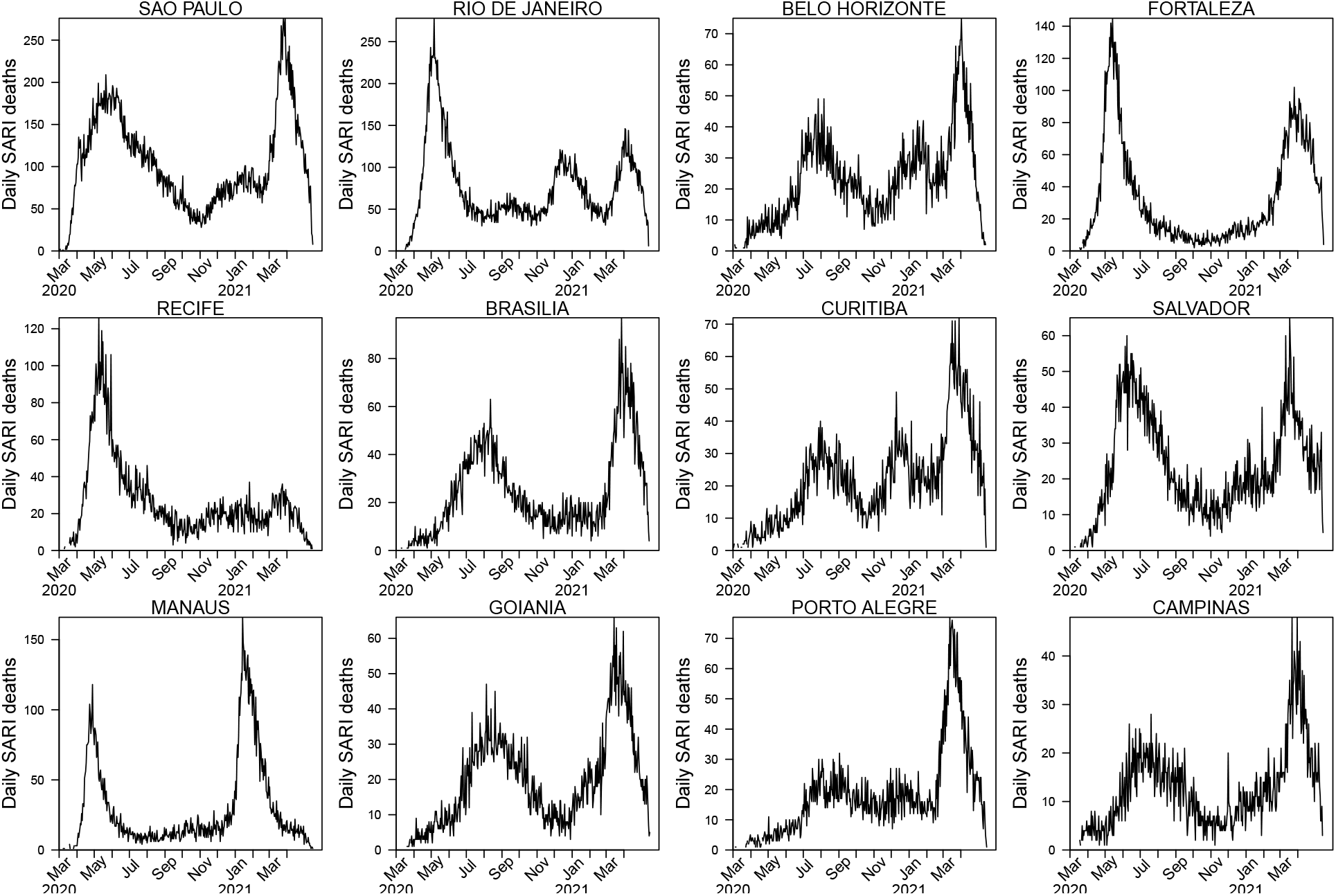
Daily SARI daily deaths in cities across Brazil plotted over 2020-2021. Data from Ref.3. The two waves in Manaus appeared in many other cities in Brazil, with some showing a more complex multi-wave character. The control of SARS-COV-2 and thus the daily deaths is a function of the cities mitigation activities, the build up over extended time of immunity, and the appearance of the highly transmissible variant of concern P.1 which spread through most of Brazil in December 2020 after spreading from Manaus.

This major misassessment of attack rate and herd immunity status, warrants further serious investigation of the epidemiological processes that took place over the first wave. Here we explore alternative and complementary epidemiological modelling analyses of publicly available mortality datasets to help understand why the first wave of the COVID-19 outbreak “crashed,”, why a second wave suddenly appeared in Manaus in December 2020 contrary to expectations, and what led the second wave to be larger than the first. We provide a parsimonious model that accurately reproduces the two-wave and two-strain COVID-19 dynamics in Manaus to resolve these contradictions, without the need to assume large reinfection rates, as in other studies (3, 5). The modelling analysis also gives important information on the attack rate (AR), the impact of non-pharmaceutical interventions (NPIs), the questionable attainment of herd immunity, and provides estimates on the transmission of the SARS-CoV-2 virus and the new P.1 lineage.

Currently there are only speculations as to why a major second wave developed and how it could have grown to such levels (4). One possibility is that immunity gained from the first epidemic had waned quickly enough to allow reinfections. An alternative possibility is that the different SARS-CoV-2 strains that dominated the first and second waves are antigenically different enough not to elicit effective cross-immunity to reinfection. But all such explanations rely on large numbers of reinfections fuelling a second wave. If we assume the original population estimates of the study are correct (AR≃76%)(3), a simple calculation shows that this would require ≃50% of the city’s 2.2 million residents to have been infected twice, once in the first wave and then again in the second. But unusually, to date there is no evidence to support these high reinfection numbers. On the contrary, as of April 10 2021, only three reinfections have been confirmed (6, 7) by local health authorities in Amazonas and only 29 in all of Brazil (by June 5 2021), out of >18 million cases COVID-19 cases. Surveillance at the authors (FN and TG) Institutes in Fiocruz in any case finds reinfection numbers similarly minimal (see Discussion). Some researchers have also suggested that the ∼76% Attack Rate estimate (AR), obtained by seroprevalence testing, may have been inflated as a result of biases (8, 9) in the blood donor data (3). Despite the difficulties in reconciling these issues with the conclusions from the seroprevalence study, a new updated analysis of the Manaus epidemic (5) still posits a mean attack rate estimate of AR≃76% up until October 2020 (see Fig.4 in Ref. (5)), indicating the same authors have not repositioned their view significantly.

The modelling approach proposed here is intended to quantify what happened in Manaus more accurately. The model is especially designed to take human behavioural responses and non-pharmaceutical interventions (NPI’s) into account when modelling the spread of COVID-19. Such mitigation activities act to depress the size of epidemics by reducing the reproductive number *R*_0_(*t*) over time (10-14) providing a “protection” effect based on behaviour rather than immunity. Despite the infamously disorganized and often ineffective interventions in Brazil, there were still significant time periods when mitigation practices had impact, and these periods need to be taken into account when making quantitative assessments. The key to modelling is finding a way to estimate *R*_0_(*t*), which is a measure of the transmission rate, as it changed over the epidemic. In more formal terms, the reproductive number *R*_0_(*t*) is defined as the average number of secondary infections a typical infectee can generate over the period of the disease, at time *t*, in a fully susceptible population. Brauer (10) modelled behavioural interference by assuming that *R*_0_(*t*) reduces exponentially over time while other studies of cholera and influenza use similar parametric approaches (11, 15). Here we generalise this principle by fitting a “flexible” *R*_0_(*t*) to the data without pre-imposing any specific functional form (12, 13, 16). Our method makes it possible to reconstruct how *R*_0_(*t*) changes in time, and from this also gain some insights the degree to which interventions impacted the epidemic, and factor this in when calculating AR and levels of immunity in the population.

### Data

The analysis makes use of two types of daily mortality datasets that are publicly available. Mortality data should reflect trends in disease dynamics with reasonable reliability and provides a useful reference frame, particularly when the quality of other surveillance datasets might be in question, as is often the case. The first dataset, provided by Brazil’s Ministry of Health (1), consisted of Severe Acute Respiratory Illness (SARI) daily deaths taken from hospitalized cases (including COVID confirmed) across Brazil. The same SARI deaths have been used in a number of recent key studies where they were considered a proxy of true COVID mortality (3, 17). The data is available for cities and all 27 confederate units in Brazil. For Manaus and Amazonas, we also analysed the COVID-confirmed mortality (CCM) datasets (18) which were continually updated with retrospective corrections. The data was collected and compiled by the local government, and in collaboration with the FVS (Fundação de Vigilância em Saúde do Amazonas) and Brazil’s Ministry of Health (18). The two datasets are compared in Fig. 1 for the two waves of the epidemic in Manaus and Amazonas, beginning in March 2020. The CCM dataset of daily deaths appears to suffer from relatively minor under-reporting, but only to a limited extent, and it is quite likely that the CCM dataset is a proper subset of the SARI deaths. When estimating attack rates, we only used the SARI dataset so as to minimise the effects of any under-estimation. In general, we repeated all analyses on the two datasets and there was generally little difference.

## Results

In order to explore the dynamics in more depth, we fitted an SEIRD-type epidemiological model to the mortality datasets. This requires compartmentalizing the population into mutually exclusive classes based on epidemiological states (i.e., Susceptible, Exposed, Infected, Hospitalized, Recovered and Dead) and modelling the disease transmission between the compartments. The model was later extended when needed, to a two age-class model and multiple-strain model. A full description can be found in the Methods and SM5.

Turning specifically to Manaus, we explored several different scenarios, which are discussed in turn.

### 1. Largely unmitigated epidemic

We began by asking the question: “Was the first wave a largely unmitigated epidemic?” This was suggested in Ref.1. A largely unmitigated epidemic would correspond to a model with a fixed transmission rate i.e., *R*_0_(*t*) = *constant*. which is, in fact, the usual configuration of SEIR-type models when simulating single epidemics. We fitted a constant transmission model to the data, but it was found that any such fit yielded outcomes that did not appear to be meaningful. For example, Fig.3a shows the best fit to the Manaus daily death data with a constant transmission rate. The modelling predicted a large fixed *R*_0_ = 3.1 over the whole period through to January 2021 (blue dashed line), and provided a very poor fit to the data. Specifically, the observed epidemic curve (of the daily deaths -- red) sit far from the median while the 95% range based on 1,000 simulations (grey shaded region) is extremely broad. Hence, we reject the possibility of Manaus having experienced a largely unmitigated epidemic in the first wave.

### 2. Modelling with flexible R_0_ (t)

We then proceeded to model the mortality data with a flexible fitting of *R*_0_(*t*) so as to estimate and allow for changes in *R*_0_(*t*) as the epidemic evolved in time. This would allow determination of when and to what degree *R*_0_(*t*) changed, presumably due to mitigation activities. Figs. 3b&c show that for this scenario an excellent fit of the Manaus mortality data is readily obtained. Fig. 3b gives fits for CCM mortality data (from Ref.(18)) while Fig.3c fits SARI mortality data. Note how the observed epidemic dynamics (red line) sit within the 95% range based on 1,000 simulations (grey shaded region) and close to the median of the model simulations (black). Moreover, the SEIR-type model, despite being acclaimed for its ability to fit epidemics of all types, was unable to do so here unless *R*_0_(*t*) (blue dashes) decreased rapidly in April. The city’s lockdown and mitigation activities were presumably responsible for this decrease, and it was typical of regions in most of Brazil as seen in Fig.4. and SI Fig.S2.

**Figure 3.**
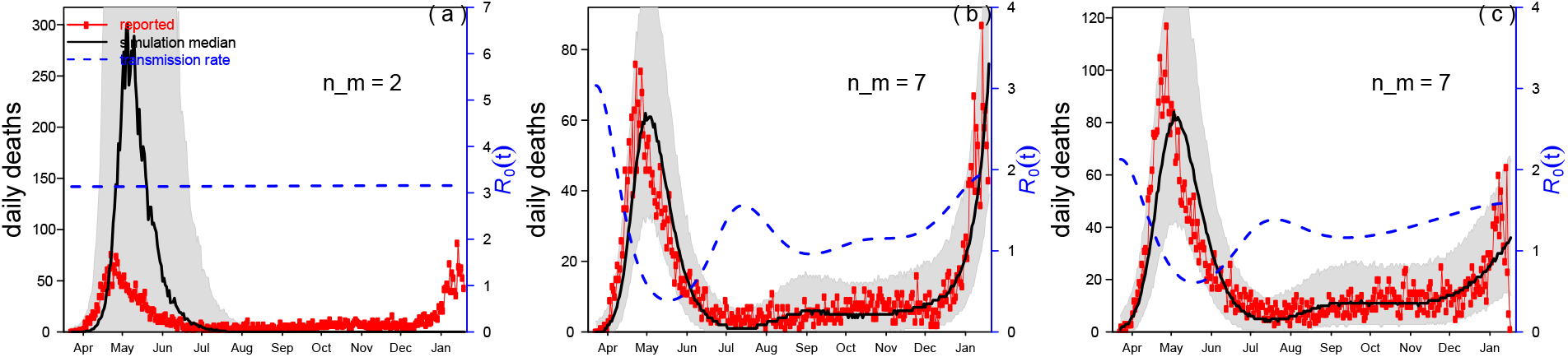
The SEIRD model was used to fit reported daily Covid-19 confirmed deaths (CCM) in Manaus (red) with a flexible transmission rate *R*_0_(*t*) (blue dashed line). The black line is the median of model simulations of the mortality data, and the grey shading indicates the 95% range based on 1,000 simulations. (a) A constant transmission rate (constant *R*_0_) fails to fit the major wave well (panel a; number of nodes in transmission rate spline nm=2). CCM data from Ref.(18). (b) When the spline’s number of nodes was increased to 7, this gave a flexible R_0_ (t), and the fitting performance was improved. Based on the 2nd-order Akaike Information Criterion (AICc), the best performance was found for nm=7. Data from Ref.(18). (c) Same as panel b but based on SARI mortality data in Manaus from(1) (red).

This feature allowed a low stationary endemic state to be maintained from July to November, as was actually the case. (Note that *R*_0_(*t*) should not be confused with the effective reproductive number *R*_*e*_ (*t*) = *R*_0_(*t*)*S*(*t*) mentioned in our Supplementary Material, which is not plotted here.) In the first phase of the epidemic, our estimated reproduction number for Manaus is *R*0 =2.250 95% CI(1.79, 2.79) (see SM1) with data from(18), which is similar to other studies (17, 19, 20). A full test of the *<u>“</u>*<u>flexible</u> *<u>R</u>*<u>_0_ (</u>*<u>t</u>*<u>)”</u> methodology is given in the Supplementary Notes (SI10).

The model estimates found that by October 2020 in Manaus, the AR≃30% which is far less than the AR≃76% in Ref.(3), and far less than the herd immunity threshold estimated as 60-66% also in Ref.(3). It would nevertheless appear to be reasonable given the large ongoing second wave that re-emerged in December only a few months later. We will discuss this further shortly.

Buss et al.(3), argued that a high estimate of AR≃76% for Manaus in the first wave, implies that the infection fatality rate (IFR) must be extremely low, in order to maintain consistency (it was claimed on average IFR≃0.018-0.26%). However, we were unable to fit mortality dynamics sensibly when assuming such a low values of IFR (see Figure S3). Buss et al. suggest that the IFR of Manaus was almost 40% lower than that of São Paulo. But this appears to contradict what is observed in the ASFR (Age SARI Fatality rate which should be a proxy for the IFR) data, as shown in SI Figure S4. In fact, the ASFR of Manaus is larger than most other cities in Brazil. This conflicting information makes the comparison between Manaus and São Paulo complex and insights regarding the differences in AR inconclusive, despite its use in other studies.

### 3. Attack rate of first wave until October 2020, also considering population demographics

In order to estimate attack rates, we also included the populations’ demographic characteristics to take into account e.g., the fact that older members of the population are more susceptible to COVID. Manaus can be qualitatively approximated as a two age-class system. Some 20% of the population are older than 45 years of age with high infection fatality rate (IFR) and contribute significantly to COVID-19 related deaths, while the remaining 80% of the population are under 45 years of age and have such low IFR that they contribute little (<7%) to the COVID deaths, according to Levin et al, (2021)(21), in an analysis of the first wave. Thus, when estimating attack rates we also made use of a two age-class model, which was constructed as a natural extension of the basic model (see Methods). Qualitative arguments (given in SI5), however, led us to believe that adding the age-class dynamics might not change the results in a major way, as was indeed the case.

The model estimates found that by October 2020 in Manaus, AR≃34% with a two age-class model, as based on SARI mortality data.

### 4. Modeling the full two-waves with a two-strain model

Following the rapid explosion of the second wave of COVID-19 in December 2020, a new highly transmissible P.1 lineage of SARS-CoV2 was identified. Initially it had been described at around January 11, 2021, but the same lineage was also later identified in samples that date back to early December 2020, most likely already present in November 2020 (5, 20, 22). We therefore attempted to also model the full two-waves of the Manaus epidemic using an explicit two-strain model (Fig.5a) that included both the P.1 and non-P.1 variants. Given the significantly low cases of re-infection found to date, it was assumed that the strains share full cross immunity, such that recovery from infection by one of the strains, precludes becoming infected by the second strain. Structurally, the second strain was integrated into our original model by adding compartments that explicitly represent the different epidemiological states associated with the P.1 strain (i.e., Exposed, Infectious, Hospitalized, Recovered and Dead).

This was also an opportunity to estimate the unknown transmission rate of the P.1 variant. As such, the parameter *η* was introduced to denote the relative transmission of P.1 variant compared to non-P.1 strain.

To facilitate the partitioning of the model fit between two strains we used data from Naveca et al.(20) that gave the relative proportion of P.1 cases among all cases as a function of time based on genomic analyses as plotted in Fig.5b (see also SI Fig.S7). The goal was to check whether both simulated daily mortality data would match observed data, and simulated strain proportion would match observed strain proportion (open circles in Fig.5b) as a function of time. The value of the parameter *η* which gave the best fit to the data was deduced numerically (i.e., in terms of the smallest sum of squared error (SSE)).

Fig. 5a shows the results of the two-strain model fit to the SARI data up until March 1 2021. In this fit, 34% of the population was infected by October 2020, while 67% was infected by the end of February 2021. To the best of our knowledge, this is the first model for which the two waves of the observed data falls within the 95%CI of the model simulations (cf. Refs (22, 23)). A good fit to the population data was only possible if the P.1 transmission rate was 1.9-fold that of the non-P.1 strain (22, 23) as seen in Figures 5b,d. This is similar to estimates in (22, 23) which suggested the transmission rate of the P.1 was 2.2 timed greater, although based on smaller sample size for their genomic data (SI Fig.S7). (Naveca et al (20) gives several hundred sequence samples per month compared to Faria et al. (5) who used a sample size of several dozen per month.) The fit of the genomic data in Figure 4b is at least as good as, and mostly better than other attempts (22, 23). Finally, it should be noted that similar to all other related studies we are aware of, the model did not take into account that during the second wave of the Manaus epidemic, the proportion of those infected in the population of younger age (also having a lower IFR) significantly increased. This would tend to increase the Attack Rate making our estimate of AR≃67% conservative.

**Figure 4.**
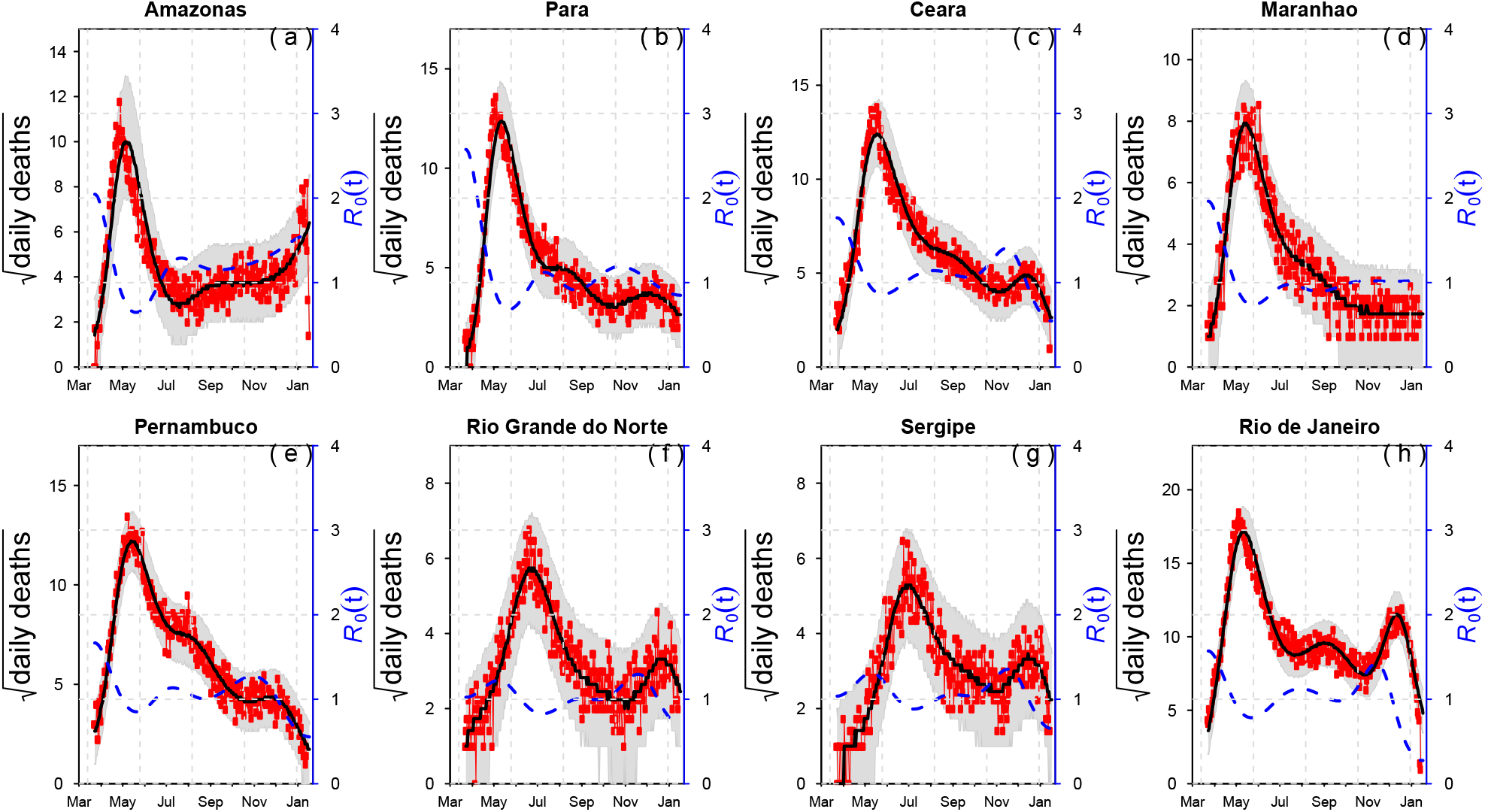
An SEIRD model is fitted to reported daily SARI deaths in eight Brazilian states (1) (red). The number of nodes in transmission rate spline was set at n_m_ =7. Red points represent real data in terms of deaths per day, the black line is the median of model simulations of the mortality data, and the grey shading indicates the 95% range based on 1,000 simulations. Supplementary Figure S2 gives plots for all other federative units

**Figure 5.**
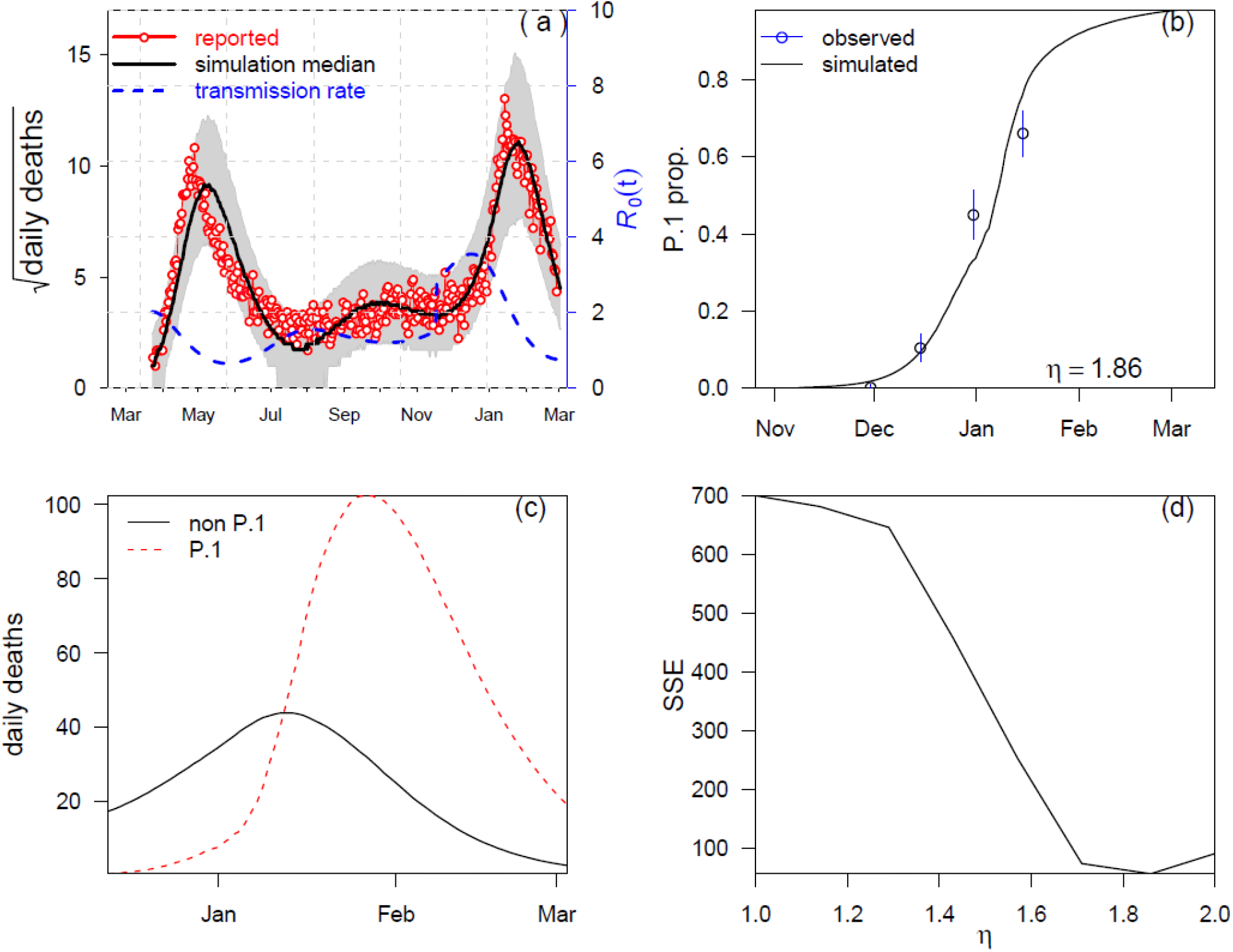
Fitting a two-strain model to SARI mortality data and sequencing proportion of P.1 strain simultaneously. *η* denotes the relative transmission of P.1 variant compared to non-P.1 strain. *η* = 1.9 yields the best fit for both SARI deaths (a) and sequence proportion (b), the latter taken from Ref.21. Panel (c) shows the simulated daily deaths due to P.1 and non-P.1 strains. Panel d shows the sum of squared error (SSE) based on sequence proportion data (b) as function of *η*. Smaller or larger *η* leads to mismatching between simulated and observed sequence proportion.

## Discussion

It is not uncommon for potentially large-scale epidemic outbreaks to die out well before predictions of conventional epidemiological models, and well before herd-immunity has developed (10). Brauer (10) explored this when studying the Ebola outbreak in Sierra Leone Guinea 2014 (24), which began with an initially susceptible population of 5,360,000 residents. While a conventional SEIR-type model predicted ∼5,000,000 Ebola cases (10), the epidemic resulted in “only” 14,120 infected individuals. We documented a similarly perplexing account of a catastrophic epidemic outbreak in WW2 (12). Using a similar modelling approach to that proposed here, it was shown that a dangerous outbreak of pandemic typhus crashed well before herd immunity was reached, and just before winter when the number of infected cases should accelerate. Brauer (10) demonstrated that the early demise of an epidemic can occur in the presence of: i) highly heterogeneous transmission and super-spreader-like events (13, 24); ii) human behavioural responses which act to depress the size of epidemics by reducing the reproductive number *R*_0_(*t*) over time (10-14), providing a “protection” effect based on behaviour rather than immunity. Here we attempt to take the latter into account by fitting a “flexible” *R*_0_(*t*) to the data (12, 13, 16).

The evolution of *R*_0_(*t*) in Manaus and Amazonas was similar to what was found right across Brazil as can be seen in Fig. 4 which plots the daily number of SARI daily and our estimates of *R*_0_(*t*) in eight arbitrarily selected Brazilian states (data from Ref.(1); Supplementary Figure S2 gives plots for all other federative units) (Again note that these are plots of *R*_0_(*t*) and not *R*_*e*_ (*t*) = *R*_0_(*t*)*S*(*t*) which is confounded with the susceptible numbers and thus a poor guide to the impact of interventions.) The graphs indicate that the epidemic curve of the first wave was roughly synchronized over all of Brazil in the period May-July. All 27 federative units peaked in this 2-month period, some earlier than others, and then declined within several months. The differences in timing between states presumably relate to spatial spread and differences in local NPI policies in terms of initiation and intensity as well as differences in local physical circumstances. The outbreaks in the first wave appear to have turned around due to the impact of mitigation procedures in Brazil (NPIs including social-distancing, facemasks, hand-sanitizing, work stoppages, lockdowns, travel bans, restrictions on gatherings, etc.) and not the attainment of herd immunity. This is seen in the major reduction in the reproductive number *R*_0_(*t*) in Figure 4 early in the first wave of the epidemic in most regions. Several studies have attempted to correlate social distancing with drops in the reproductive number in Manaus (3, 20). The epidemics curtailed even though the mitigations activities were often flawed, mishandled, and often not taken seriously by many sectors of society including powerful figures in government (see SI8). Better handling of the public health catastrophe could have prevented the extreme death rates seen in Manaus, and all over Brazil.

The model estimates an attack rate of AR≃30%-34% up until October 2020, and thus far below herd immunity. This is almost identical to results found in a very recent independent seroprevalence study based on 3046 residents of Manaus (25). For the same period Lalwlani et al. (25) estimated a crude AR of 29% (but possibly up to an adjusted AR of 40% when using an atypically high seroreversion rate). Our estimates of AR are also in-line with the work of Mellan et al. (19) and Hallal et al. (2) who reported similarly low attack rates. The seroprevalence survey of Hallal et al. (2) reported an AR=14.6% with 95% confidence interval CI: (8.9%, 22.1%) by June 7, 2020, while our model estimates AR= 20% by this time.

In the process of calculating the AR, our model takes into account dynamical features of the epidemic trajectory over time. The evolving shape of the trajectory itself is a source of information on the underlying dynamics and provides a more sophisticated assessment relative to gross calculations based, for example, on IFR estimates and seroprevalence data, particularly in cases where the latter may potentially be prone too sampling biases.

### Reinfections

Although the results reported here quantitatively confirm that the AR in Manaus was likely to be far less than 76% by October 2020 (as estimated in Ref.(3)), they point to another danger that was overlooked at the time – the possible development of a second wave. With the appearance of the second wave in December 2020, it has become clear that either the estimated (3) AR of 76% was incorrect, or there were widespread reinfections generating the second wave. But to date only some 3 confirmed reinfections have been recognized by the health officials in Amazonas (7), so this latter argument appears to be implausible. The work of the authors (FN and TP) at Fiocruz (Oswaldo Cruz Foundation), in Manaus corroborate this. All workers and theirs’ families at the Institute were tested since the beginning of the pandemic with serological tests (IgM and IgG) and PCR if they were either symptomatic or contacts of positive cases. Most cases of Fiocruz workers in the second wave were found among those that were never positive before. There were only two cases of reinfection among this group of approximately 300 persons. Another large-scale study of N=154,000 infected individuals in Israel found a reinfection rate of 0.1% (two PCR-positive at least 100 days apart)(26) again highlighting our point. Another large-scale study in Denmark found a rate of 0·65% two PCR-positive at least 3 months apart) (27). As far as is currently known reinfections are minimal in other countries. In a systematic review of 11 large-scale cohort studies in different countries reinfection of SARS-COV-2 was found to be rare (0%–1.1%) and immunity did not wane for at least 10 months post-infection (28).

The discrepancy between our model and that of Faria et al. 2021 and Buss et al will be outlined elsewhere but relate to their unusually high estimates of seroreversion decay rates, and their low estimates for IFR. The attack rate estimates of Faria et al. (5) are based on accepting an IFR that is close to IFR=0.32%, but it is hard to find proper justification. Older studies that might suggest a low IFR (21) need updating based on new data now available and in light of our calculation given above. The IFR=0.32% (used as a prior) would automatically set AR>67% in the first wave given that the records show 4,600 SARI deaths in Manaus in that period. For the case of almost full cross immunity (few reinfections), the model of Faria et al. (2021) is unable to fit the data unless the p.1 strain has an IFR three times that of the non p.1 strain, which we believe to be extreme, and largely an outcome result of trying to maintain a large AR>67% in the first wave.

The more likely explanation for the second wave is that a large pool of susceptibles still remained after the first. Thus relaxation of mitigation efforts in late 2020 combined with the appearance of the highly transmissible P.1 variant led to the triggering of the second wave, and the large numbers of deaths that rapidly followed. (The model in fact estimates that the P.1 variant is 1.9 times as transmissible as the original strain.) Moreover, we cannot rule out that belief in the “herd immunity” status contributed, at least in part, to the second wave in Manaus, by creating a false atmosphere of normality which encouraged relaxation of NPI’s and led people to expose themselves to the virus. Through modelling, we estimate that ∼70% of the Manaus population became infected by March 2021.

Our work highlights the difficulties policy makers face when attempting to predict herd immunity, as well as the importance of developing and using complementary mechanistic epidemiological modelling tools when assessing outbreaks with one or more virus strains. The methods used, here based on a “flexible number *R*_0_(*t*),” are particularly useful in cases where NPIs and human behaviour impacts the temporal development of epidemiological dynamics, for which other traditional methods can fail. The experiences in Manaus with regard to COVID-19 make clear the need for future research characterizing the nature of reinfection and protective immunity, for which we currently have limited understanding, and the epidemiological dynamics they induce at the level of the host population. Incorrect assumptions or viewpoints about underlying processes, can lead to major misconceptions on how to most responsibly address upcoming challenges of COVID-19. This will be all the more problematical with respect to characterizing the emergence of de-novo strains, and their potential trajectories of adaptation, such as immune escape, evolution of vaccine resistance and virulence.

## Methods

### The SEIHDR epidemic model

We use an susceptible-exposed-infectious-recovered (SEIR) type model with a flexible time-varying transmission rate *β*(*t*) to the reported mortality data:

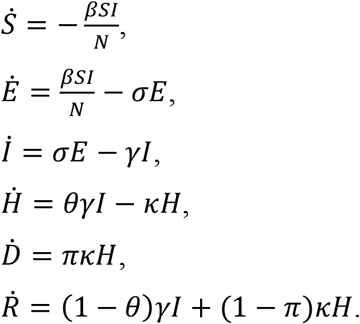

Here the compartments *S, E, I*, and *R* denote the conventional susceptible, exposed, infectious, and recovered individuals, respectively. The constant *N* is the total population size of the city or state, *H* denotes the hospitalized cases (or severe cases), and *D* denotes the total of SARI-deaths. Parameters *β*(*t*), *σ, γ, κ* denote the transmission rate, the infectiousness emergence rate, the infectiousness disappearance rate, and the removal rate (due to death or recovery) of hospitalized cases, respectively. Parameters *θ* and *π* denote the ratio of hospitalized cases out of all infected cases and the proportion of deaths out of hospitalized cases, respectively. Thus, the overall case fatality rate (or infection fatality rate) equals *θπ*. All parameters are constant except *β*(*t*) being time-varying. The above model was used for studying the Amazonas SARI mortality data(1) and a very similar model was used to study Manaus COVID confirmed mortality data (18).

We follow closely our previous work for fitting a “flexible” time varying transmission rate *β*(*t*) (12, 13, 29) based on fitting the mortality data. The former requires defining *β*(*t*) = exp (*cubic_spline*) as an exponential cubic spline with *n*_*m*_ nodes evenly distributed over the study period. After this was fitted, we estimated 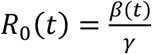. For the model, we set time step size as one day and integrated 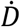 for one day to obtain the simulated daily deaths *D*_*t*_. We defined the reported deaths as *C*_*t*_, where:

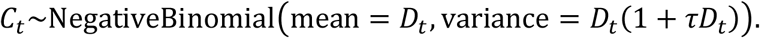

Here, *τ* denotes the overdispersion, and accounts for the measurement noise due to surveillance and heterogeneity among individuals.

The parameter values of *σ, γ, κ* were taken as 365/2, 365/3.5, and 365/12 per year, respectively. Thus, the mean latent period and mean infectious period (the reciprocals of *σ* and *γ*) were set as 2 and 3.5 days, respectively. The generation time GI,(30) the sum of mean latent period and mean infectious period, equals 5.5 days, which is in line with three key studies which provided estimated GI.(31-33) According to,(34) the delay between the first symptom onset and the death is ∼14 days, which justifies the choice of 12 days (the reciprocal of *κ*) from loss of infectiousness to death. Namely the onset of infectiousness was taken to be 2 days ahead of first symptom onset. In addition, we assume 5% proportion of the population has pre-existing immunity (35, 36), which only changes the initially large available susceptible pool by a small amount.

The Euler-multinomial simulation approach was used to simulate our model with a time-step size 1 day. The popular iterated filtering method(13, 15, 16) (note that at least 25 applications use this method, see a list at https://en.wikipedia.org/wiki/Iterated_filtering) was implemented to fit our model to the observed data and to obtain the maximum likelihood estimates of unknown parameters, including values of nodes in *β*(*t*) and *π* while holding *θ*=0.1. Namely, we assumed 10% of cases were hospitalized (or severe cases), as widely reported. The fitting is insensitive to the choice of *θ*, but sensitive to the product of *π* and of *θ*, which is equivalent to the infection fatality rate (IFR). We assumed uniform prior for all parameters. We hypothesized that the IFR lay in the interval (0.002, 0.008), and fit the daily SARI mortality data with a flexible transmission rate. The method finds the best fitting parameters including IFR. We used the 2^nd^-order Akaike Information Criterion (AICc) (12, 13, 29) to find the best number of nodes in the transmission spline *n*_*m*_. The same method was used to fit the two-age-class model.

A test of the Method on reconstructing epidemic dynamics from mortality timeseries is given in SI10.

### Age-class model

In the two-age-class model, the population in each city was divided into two age groups: <50 and 50+ years of age. The population of each group was then divided into six S-E-I-H-R-D sub-groups. It was assumed that the cross-group transmission is the same as the in-group transmission. We obtained population size and age structure (the population ratio between <50 and +50) from online sources, e.g., https://www.citypopulation.de/en/brazil/regiaosudeste/admin/são_paulo/3550308__são_paulo/ https://www.citypopulation.de/en/brazil/regiaonorte/admin/amazonas/1302603__manaus/

The daily Severe Acute Respiratory Syndrome (SARI) deaths was aggregated into two age groups: <50 and 50+ years of age, and time series were constructed for each group. The daily deaths from our model to the two time-series were fitted simultaneously. The total log likelihood of the model was taken to be the sum of the two log likelihoods for each time series.

The infection fatality ratio (IFR) was taken to be different for the two groups, with the IFR for <50 group being much smaller than that of the 50+ group. Both IFRs were estimated by our procedure. It was only assumed that the IFR for the 50+ group was between 1% to 4.5%. We assumed a flexible transmission rate (exp-cubic-spline with 7 nodes). Parameters were estimated including IFRs for both age groups, number of nodes in the transmission rates, and initial conditions. For Manaus, Figure S5 shows the fitting performance in terms of log likelihood of the two-age-class model given the data, as a function of the IFR for the 50+ group and that for the single-age-model.

## Data Availability

All data used in this work are publicly available.

## Declarations

Ethical approval and consent to participate are not applicable.

## Consent for publication

Not applicable.

## Availability of data and materials

All data used are publicly available.

## Code availability

The codes are available upon request to the corresponding author.

## Author contributions

LS, DH, YA designed the study, carried out the statistical analysis and wrote the manuscript. SSM, TG, FN, and PH critically revised the manuscript. All authors approved the submission.

## Supplementary Information for

### Section 1: Initial Reproductive number *R*_0_(*t*)

For each dataset in Fig. S1 and in the main text we plotted 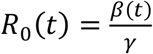, which is the dashed blue line seen in figures as a function of time (/date). The reproductive number *R*_0_(*t*) should not be confused with the effective reproductive number *R*_*e*_ (*t*) = *R*_0_(*t*)*S*(*t*) mentioned in the main text, which we do not plot. The plot of *R*_0_(*t*) allows us to visualise how the reproductive number changes e.g., as a result of behaviour, or interventions.

For Amazonas and Manaus, we also arrived at estimates for the initial reproductive number in the first phase of the epidemic, using the standard technique of Wallinga and Lipsitch.(1) This is shown in Figure S1 which is based on the COVID-19 mortality data from Amazonas government health department reports.(2)

In Figure S1, we used a standard approach, 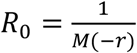, where *M* is the moment generating function of the distribution of the generation time which attains a Gamma distribution with a mean of 5.5 days and s.d. 3 days.(3, 4) We used the interval from March 30, 2020 (when there were 2 deaths) to April 12, 2020.

**Figure S1.**
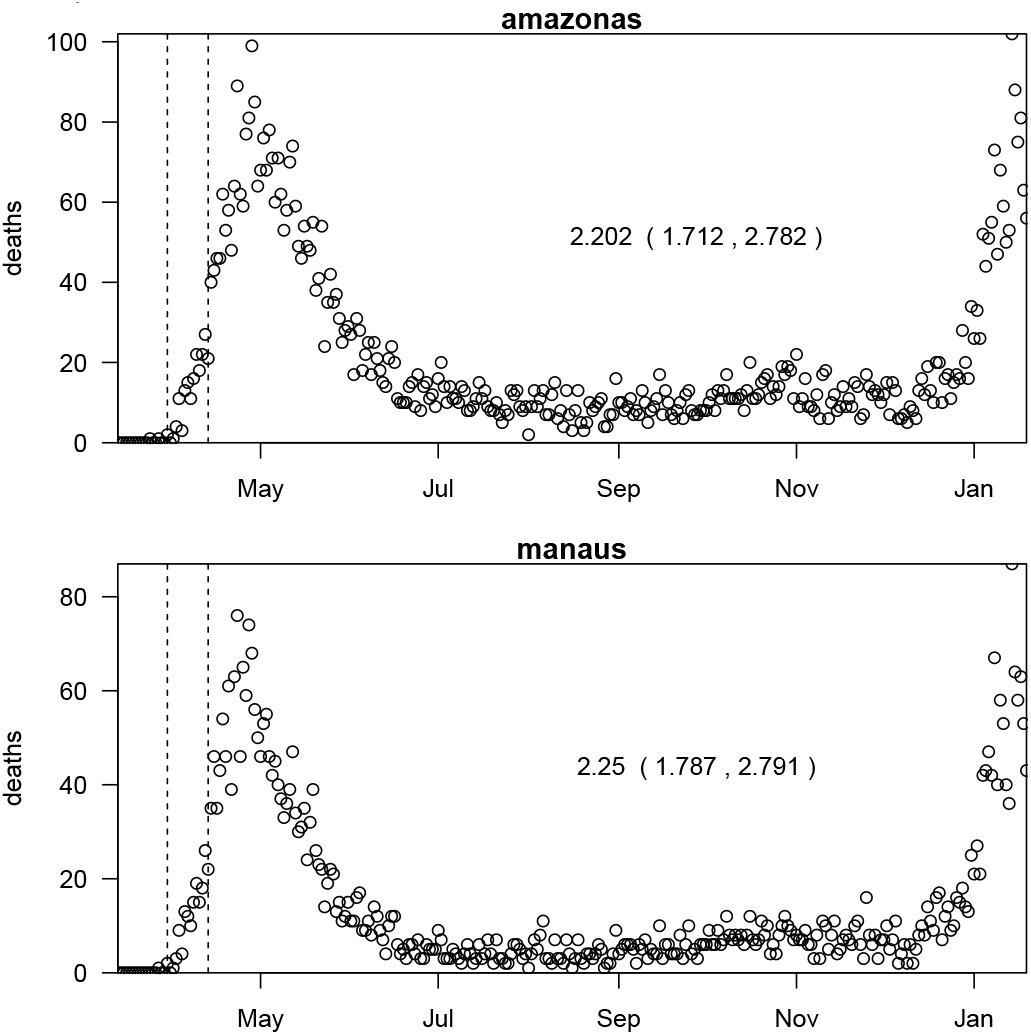
Estimating initial *R*_0_ via the method of Wallinga and Lipsitch (2007)^1^. The dashed vertical lines indicate the window in which *R*_0_ was estimated. Initial R0 in Amazonas is 2.20 with 95%CI (1.71, 2.78). Initial *R*_0_(*t*) in Manaus is about 2.25 with 95% CI (1.79, 2.79), which are in line with (5). Data are from ^2^.

### Section 2: Fitting the model to SARI mortality data in all Brazilian states

In Figure 2 of the main text, we fit the above model to SARI mortality data in eight Brazilian states. Figure S2 below repeats this for the other 16 Brazilian states and the federal district.

**Figure S2.**
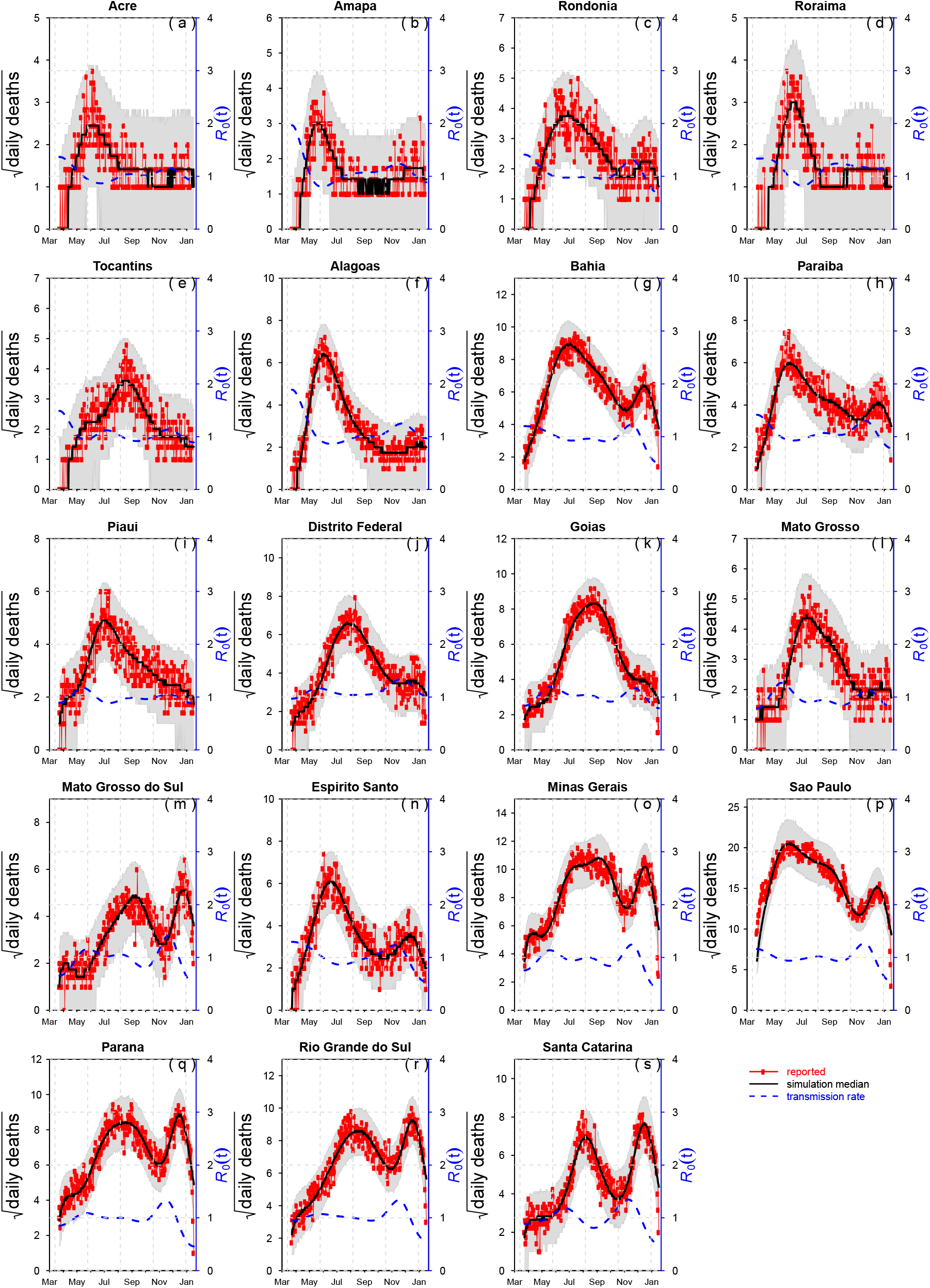
Fitting SEIRD model to daily SARI death in 16 Brazilian states and the federal district.

### Section 3: Fitting data with IFR<0.3% and Attack Rates greater or equal to AR=76% by Oct 31, 2020

In Figure S3, we explored simulations with IFR<0.3 ensuring that the Attack Rate (AR) reach levels of AR >76%, based on deaths and population size in Manaus. In all cases this assumption led to *R*_0_(*t*) being unrealistically large (i.e., *R*_0_(*t*) > 7) in the second wave. If *R*_0_(*t*)was fixed to a constant, the fit to the data was poor (Figure S3a), with the best fitting *R*_0_(*t*) reaching values of *R*_0_(*t*) > 5.

**Figure S3.**
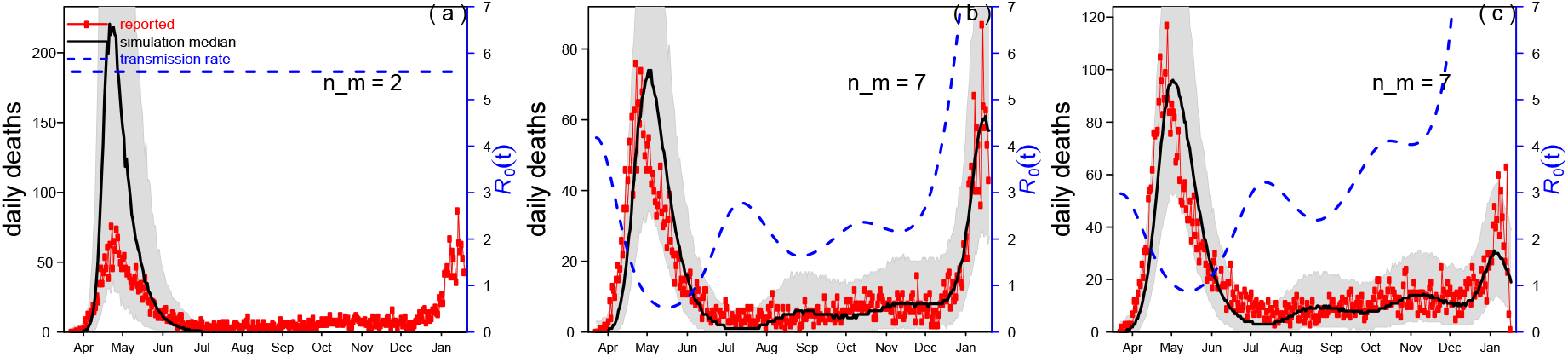
Reproducing the fitting in Figure 2 with IFR <0.3%. The estimated *R*_0_(*t*) reaches unreasonably high values during the second wave. (a,b) confirmed COVID-19 deaths. (c) SARI deaths, in Manaus.

### Section 4: IFR and AR in Manaus versus São Paulo

Buss et al. argued that the AR in Manaus (76%) was far larger than that of São Paulo (29%). They claimed this was due to the much lower global population infection fatality rate (IFR) in Manaus, an outcome of the age-class distribution of the IFR in a city with relatively younger population. They found the IFR in Manaus was 38.9% smaller than that of São Paulo. However, this difference between Manaus and São Paulo is questionable since it is not seen when examining the Age SARI fatality rate (ASFR), i.e., the ratio of deaths among reported SARI cases. In Figure S4 we plot the ASFR for four cities in Brazil including São Paulo, and find the ASFR is higher in Manaus in both waves compared to any of the other three cities. In the first wave it is particularly larger than the ASFR of São Paulo for all ages. This was also evident in plots of Buss et al. (6) in their SI.

Given the “inverse nature” of the calculations used in working with the IFR, if Manaus actually had a higher IFR than São Paulo as Figure S4 indicates, this should translate into a lower number of infected residents [for similar mortality numbers] and lower AR in Manaus, indicating another inconsistency with the suggested AR of 76%.

Also Figure S4 shows that although the population in Manaus is relatively young, we see that those of younger ages that were diagnosed as SARI, were more likely to perish in Manaus relative to São Paulo. In Manaus, the young population presumably suffered more due to the heavy burden of the epidemic conditions and the collapse of the hospital system.

**Figure S4.**
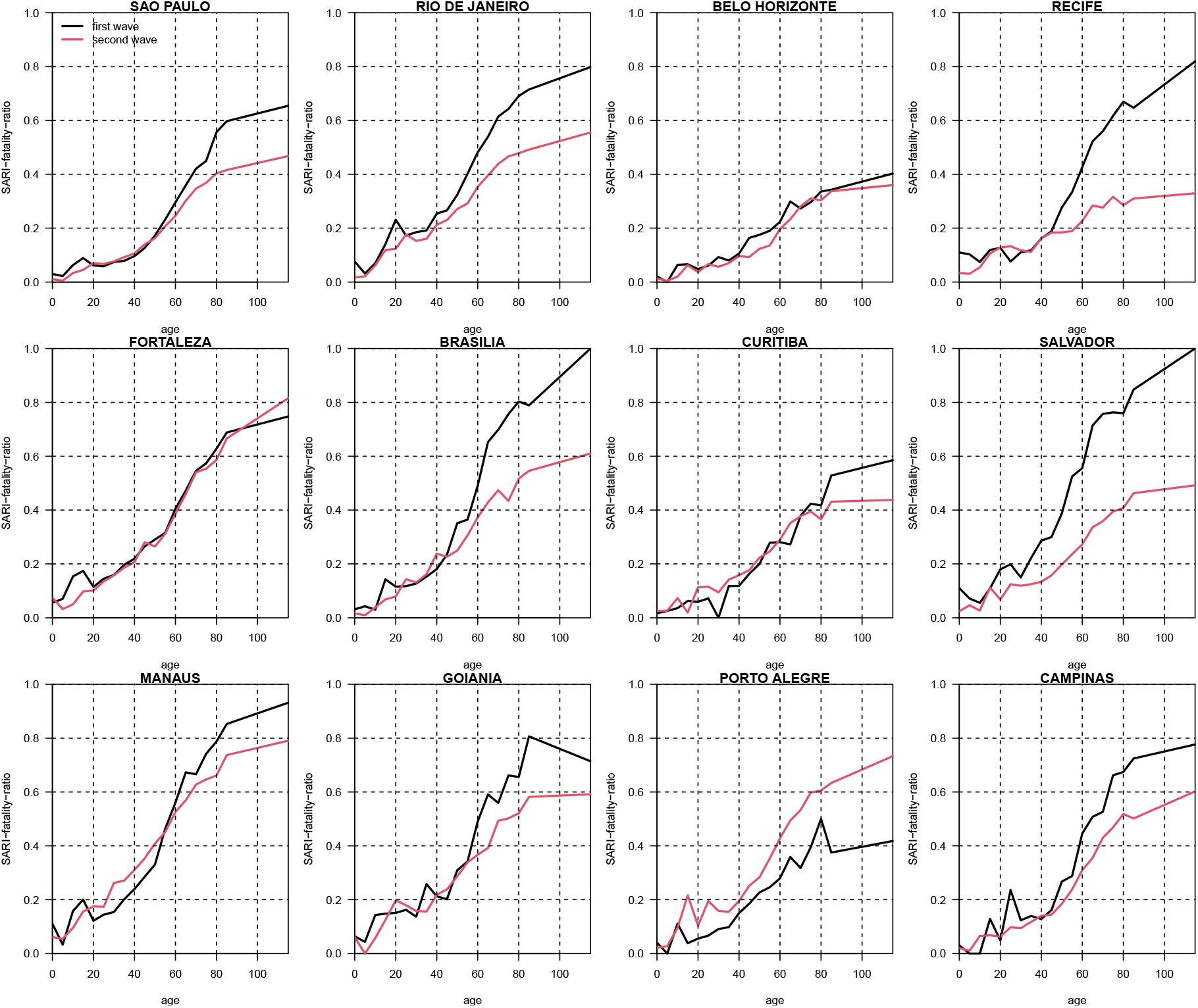
Age SARI fatality ratio (ASFR), i.e., the ratio of deaths among reported SARI cases. Black curve shows the ASFR for the first wave (March 13 - June 1, 2020). Red curve shows the ASFR for the second wave (Nov 23, 2020 – April 26, 2021). Data from Ref. (7).

### Section 5: Fitting Two-age-group model to SARI deaths including both waves

#### Age-class model rationale

Our initial results were obtained from analysing an epidemic model with only a single age-class. While this ignores important considerations of the age distribution, it nevertheless has relevance for two reasons: a) The important *Science* paper of Earn et al.(8) demonstrated that significant changes in *R*_0_(*t*) have much more impact on the epidemic trajectory than age-structure and should be the first consideration, while the effects of age-structure are in comparison far smaller. b) Qualitatively, Manaus can be characterized as a two age-class system, with people aged 45 and up significantly contributing to COVID-19 related deaths (these account for roughly 20% of the population), while the rest of the population (< 45 years) display an exceptionally low IFR, contributing very little to the total deaths toll even though the account for roughly 80% of the population (e.g., according to Levin et al. (2021)(9), they are responsible for <7% of total COVID-19 deaths. We justified our approach based on the knowledge that in contexts similar to the one dealt with here, a single age-class system found from averaging two age-classes (themselves homogenous in transmission) should be able to characterise the epidemic dynamics of the two age-class system (see e.g., Magpantay et al. (10)). Nevertheless, as a check on our work, we repeated all analyses on a complete two age-class model as described later. The results were always found to be very similar to the single age-class model, as we point out in the main text and below.

The infection fatality ratio (IFR) was taken to be different for the two groups. In particular, the IFR for <50 group was much smaller than that of the 50+ group. Both IFRs were estimated by our procedure. It was only assumed that the IFR for the 50+ group was between 1% to 4.5%. We assumed a flexible transmission rate (exp-cubic-spline with 7 nodes). Parameters were estimated including IFRs for both age groups, number of nodes in the transmission rates, and initial conditions. For Manaus, Figure S5 shows the fitting performance in terms of log likelihood of the two-age-class model given the data, as a function of the IFR for the 50+ group and that for the single-age-model.

**Figure S5.**
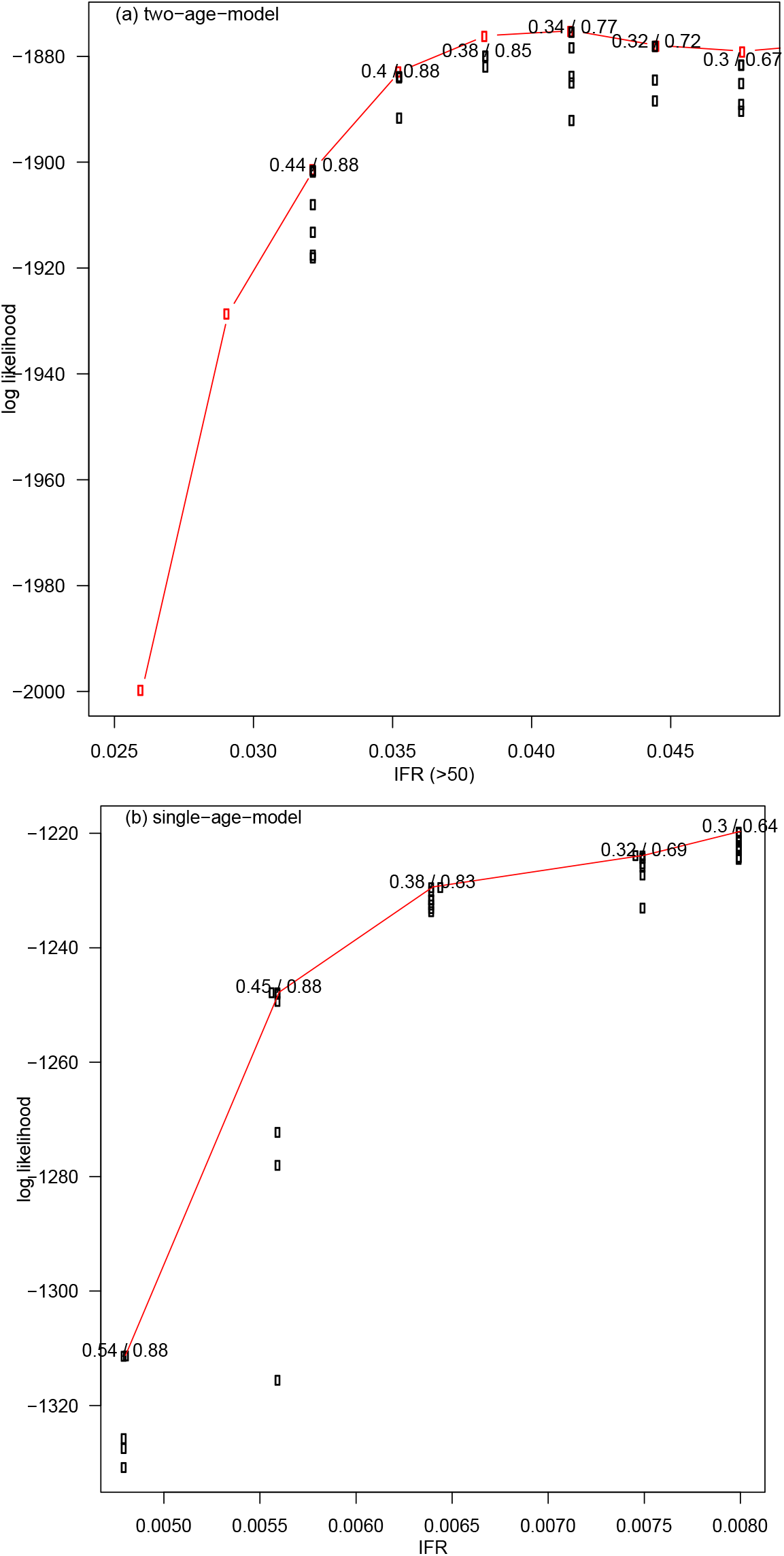
The fitting performance is plotted in terms of the model’s log likelihood given the SARI deaths as a function of the IFR for the 50+ group in (a) and of the IFR for the single-age-model in (b) in Manaus. Larger Log likelihood implies a better fit. For each of the six IFR values plotted, we give as a pair of values the infection attack rate by Oct 31, 2020 and Feb 28, 2021. The maximum likelihood estimates of infection attack rate is thus 34% and 77% with IFR for 50+ at 4.15%. Since the peak of the Log likelihood profile is very flat, 32%/72% to 38%/85% are also plausible.

We fit the model to data from 8 Brazilian cities and achieve reasonable fitting for each of the two age-classes as indicated in Figure S6.

**Figure S6.**
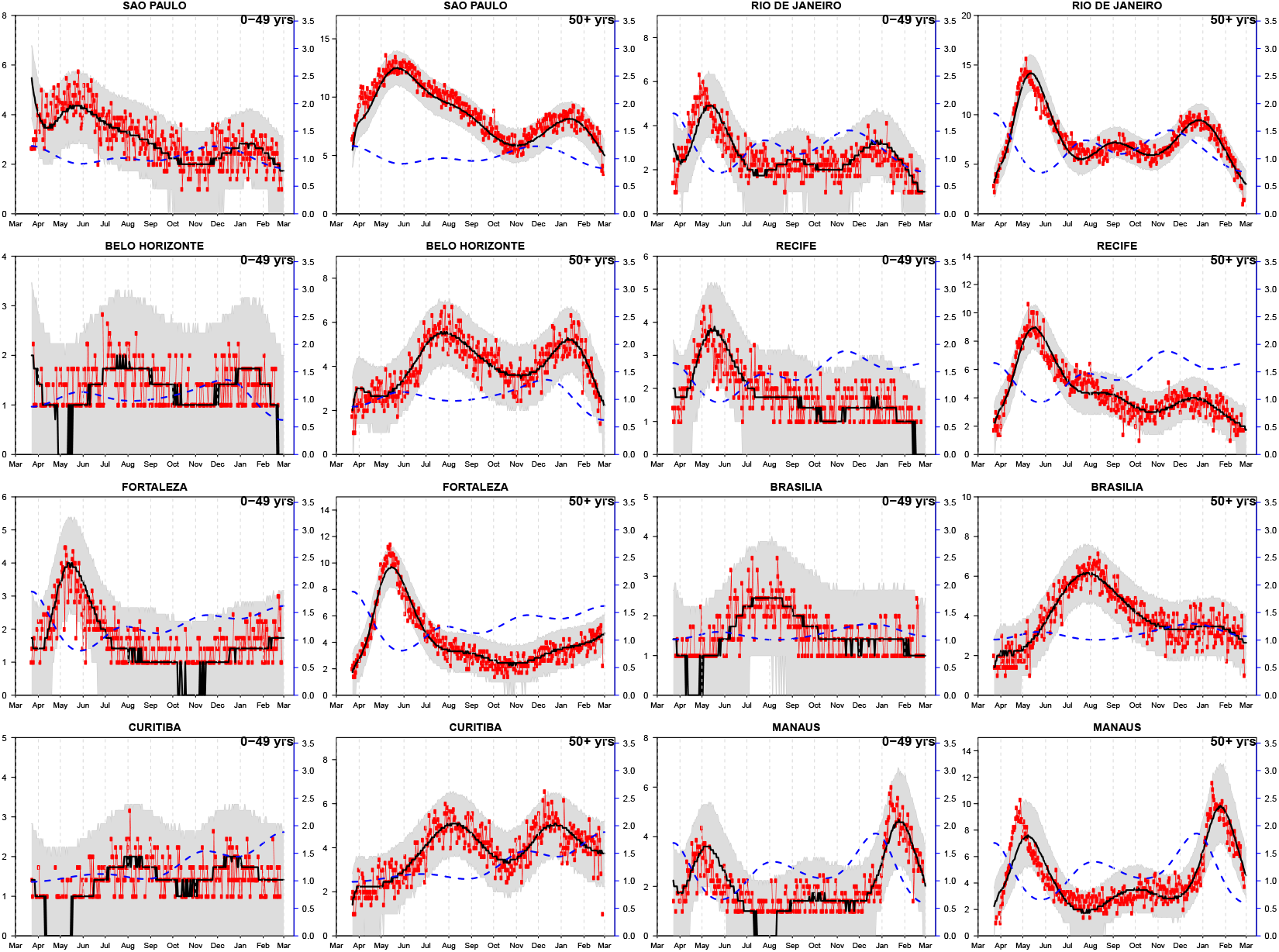
Fitting a two-age-group model to SARI mortality from eight Brazilian cities each divided into two age-classes.

### Section 6: Reinfections

If the attack rate is AR=76% up until October 2020 and let us assume that the first wave is roughly as large as the second wave. Then a simple Venn diagram would show that one needs >50% re-infection rate among infections in the second wave in Manaus. Yet currently there is no evidence or reliable reports that indicate the reinfection rates were more than 1 percent in the second wave December 2020 -February 2021. This would appear unusual if reinfections were actually occurring at high rates. The Manaus team has discussed low reinfection rates at Fiocruz Manaus (see Discussion), A large-scale study in Israel found a rate of 0.1% (two PCR-positive at least 100 days apart) (11). The other large-scale study in Denmark found a rate of 0·65% two PCR-positive at least 3 months apart). (12) Only 3 reinfections were found in Amazonas (see main text).

### Section 7: Blood-donor data

It is difficult to explain the high estimate of the attack rate of COVID-19 in Manaus, as found by Buss et al. (2021). One possibility are the sources of bias that might be introduced from working with blood-donor data. These are not discussed in their article, and thus difficult to comment on. For example, a recent COVID seroprevalence study in Kenya (13) made use of “family replacement donors (FRDs) who provide a unit of blood in compensation for a transfusion received by a sick relative.” This may have led to major biases in the Kenyan analysis. Replacement blood donation is a common practice in middle and low-income countries. According to the Agência Nacional de Vigilância Sanitária - Anvisa (2019)(14) report of Brazil, 54.3% of blood donors are repeated donors, and 37.3% of blood donors are family replacement donors in the North region. The Amazonas region may have an even higher rate of replacement donations. Screening measures (“prospective donors could not have had flulike symptoms within the 30 days before donation; had close contact with suspected or confirmed covid-19 cases in the 30 days before donation”)(15) could prevent donation from recent symptomatic cases, but not from asymptomatic cases and past symptomatic cases (who showed symptoms beyond previous 30 days). Other biased sampling of the Manaus population was also possible. For example, during the peak of the pandemic, blood donation experienced shortage, and military members were recruited(16) in Amazonas. Interviews in the media report other potential sources of bias such as donors being more mobile, or receiving free COVID tests or being a nonrepresentative sample of the population(17, 18). The study of Buss et al., could possibly be free of all such biases, but further clarification is needed.

### Section 8: Interventions in State of Amazonas, Brazil

It is worthwhile summarising that mitigation practices occurred over the whole state of Amazonas. From March 2020 to May 2020, most economic activities were closed, returning at the begging of July 2020. In late December 2020, new restrictive measures were adopted due to the rapid increase in COVID-19 cases. Since February 2021, a gradual return to normality started.

Further details may be accessed in: http://www.pge.am.gov.br/legislacao-covid-19/ (Portuguese)

### Section 9: P.1 new variant percentage among all samples sequenced

**Figure S7.**
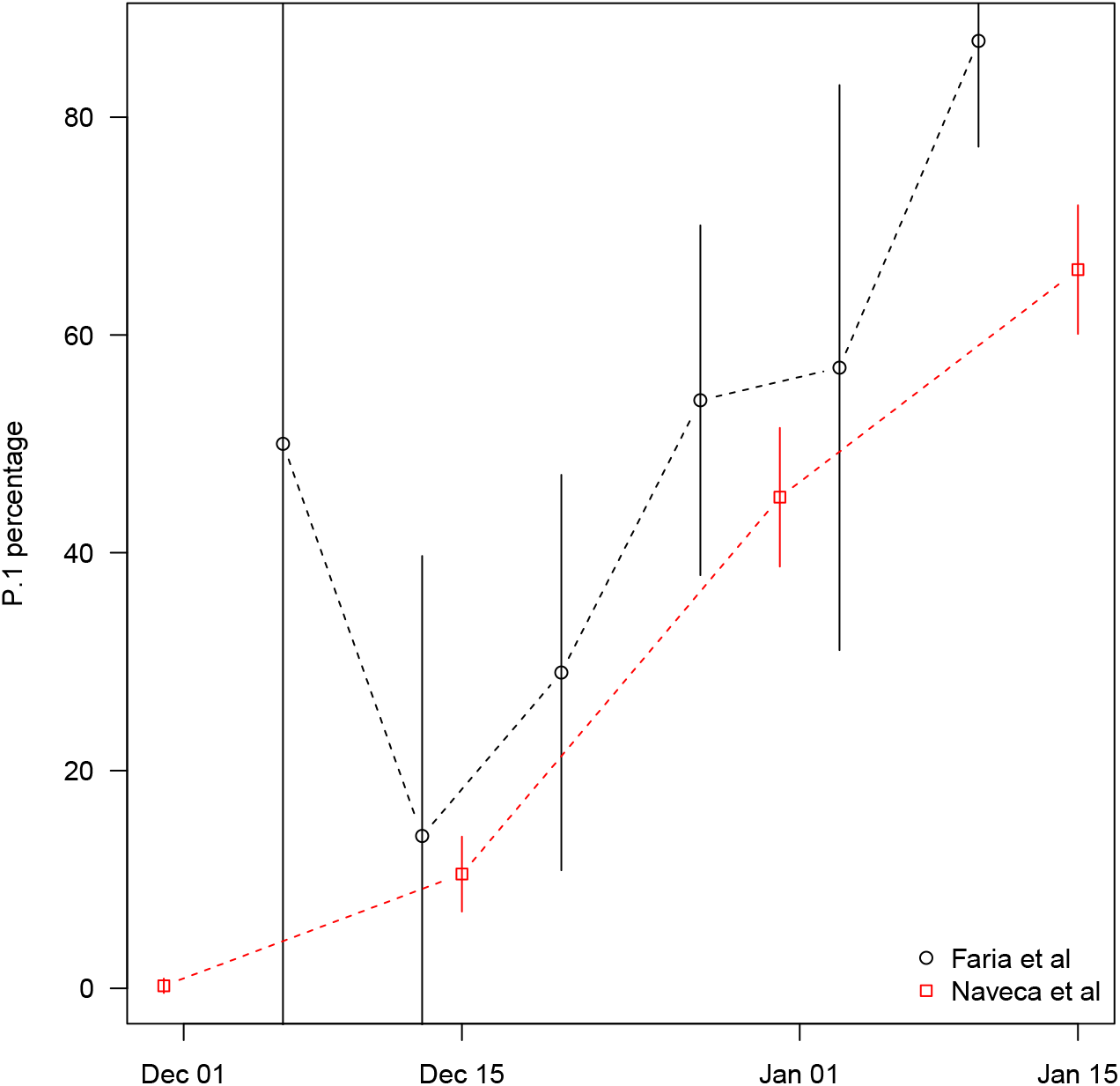
A comparison of P.1 new strain percentage among all samples sequenced between two works. Naveca et al(19) has a larger sample size of several hundreds per month compared to Faria et al(20) who had sample size of several dozen per month.

### Section 10: Checking the method

Finally, as a test of the methodology, which is Bayesian based on iterated filtering (21), we generated three fictitious time-series from March 2020 to January 2021 with *R*_0_(*t*) arbitrarily varying in time 0.5 < *R*_0_(*t*) < 2.2 (Figure A1a,b&c, blue dashed curve). These time-series were used to drive the model parameterised for Manaus in order to generate dummy datasets of daily mortality numbers (red trajectory with circles). We could then use our method to reconstruct *R*_0_(*t*) (plotted in green). The results show that the reconstructed *R*_0_(*t*) is a very close estimate of the original value (dashed blue) in each case. Similar to Figs 3-5, the black curve and grey regions, were based on simulations of the SEIRD model using the same reconstructed *R*_0_(*t*). This confirms the capability of the method to reconstruct *R*_0_(*t*) and that our analysis of the Manaus data is reliable. Variations of the methodology has been tested in other contexts in our own work and by several independent teams (22-24).

**Figure S8.**
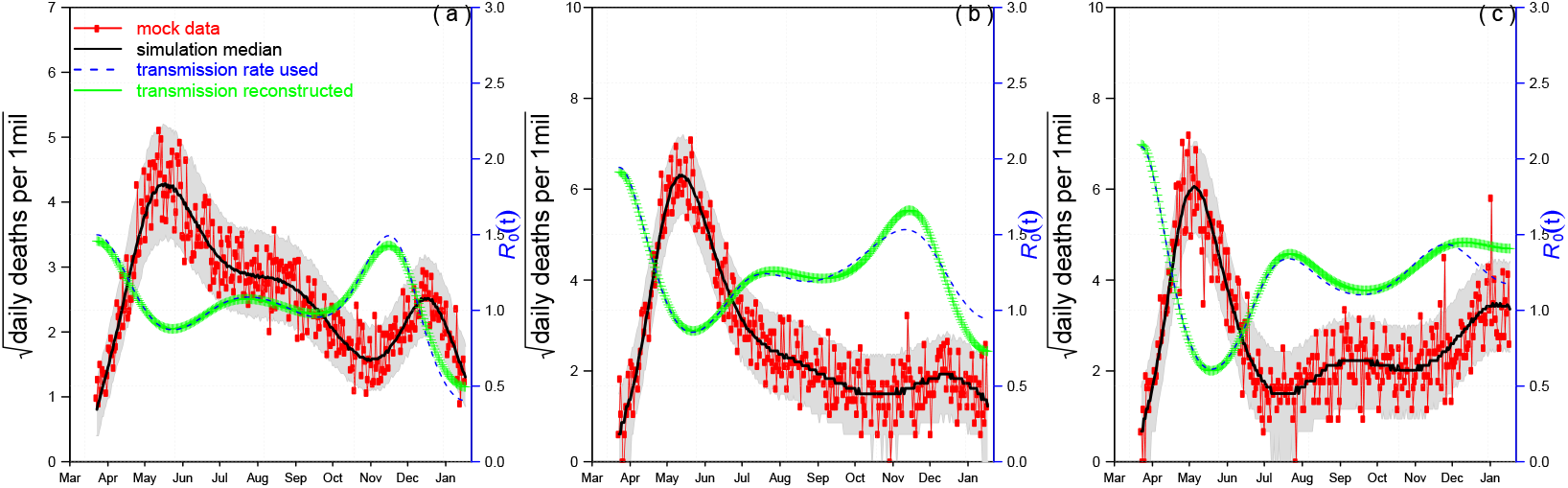
Fitting *R*_0_(*t*) from mock data. We generated three simulated time series of *R*_0_(*t*) changing over time in an arbitrary way between March 2020 and February 2021 (blue dashed lines). Each time series of *R*_0_(*t*) was used to generate a simulation of mortality data from the SEIRD model (mock data, red trajectory with circles), and basically three different patterns of death data resulted as seen in (a), (b), (c). Then we used our fitting method to reconstruct *R*_0_(*t*) (green trajectories) from the mock data. The solid black lines and grey regions, similar to Figs 1&2, were based on simulations using the reconstructed *R*_0_(*t*). The reconstructed *R*_0_(*t*) proves to be a good estimate of the initial *R*_0_(*t*).

### Section 11: Comments about the similar IFR of of P.1 and non P.1 variants of SARS-CoV-2

**Figure S9.**
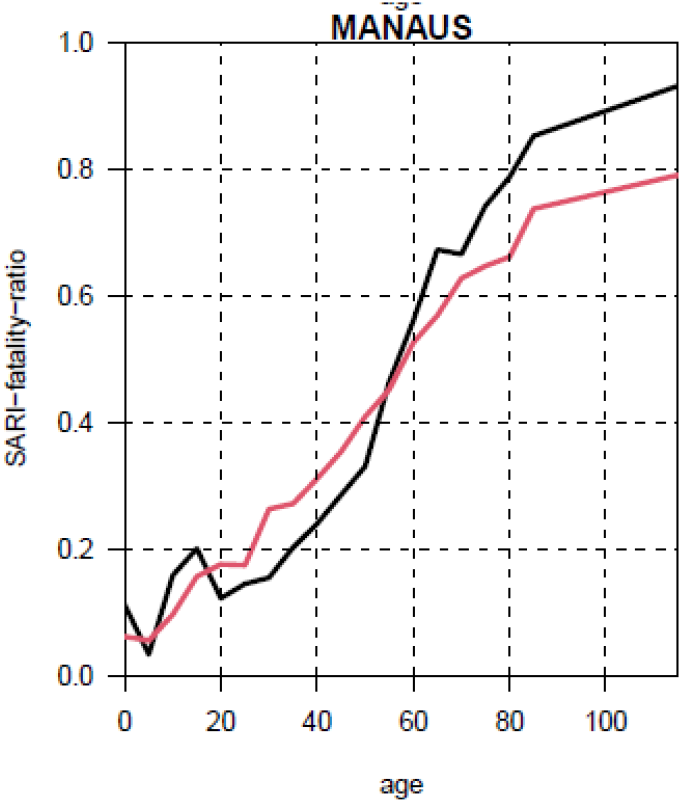
SARI-fatality ratio of deaths to hospitalised cases. First wave ratio plotted in black; second wave ratio plotted in red. Because the second wave was predominantly P.1, we can say that the lethality of the two strains, P.1 and non-P.1 (first wave) is reasonably similar. The average SARI-Fatality Ratios prove to be similar.

**Figure S10.**
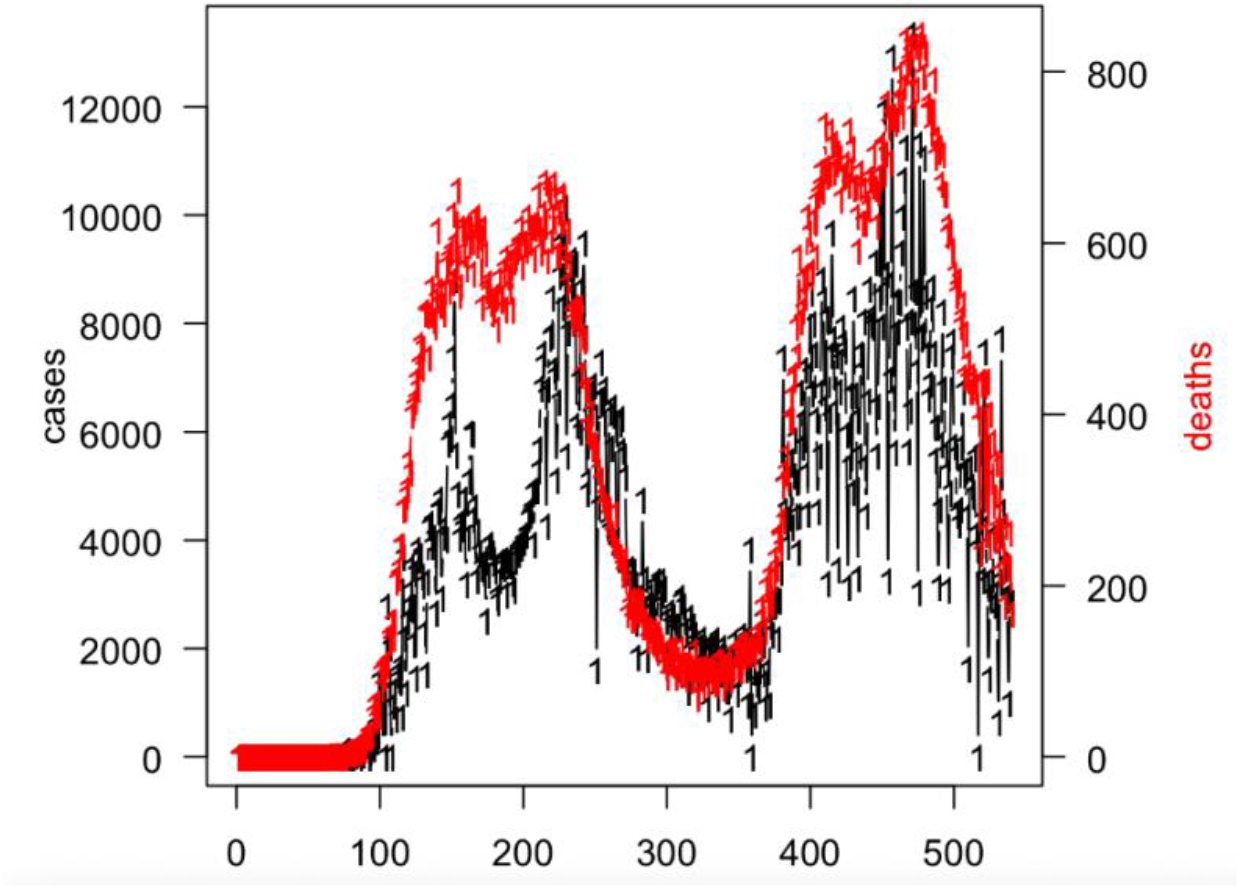
Data of confirmed COVID cases and deaths in Peru over the two waves in 2020/2021

Data from: https://covid19.who.int/info/. Note that the raw case fatality ratio is similar over the two waves, and if anything decreased in the second wave. Although changes in testing practices may have great influence, nevertheless there is little evidence from this data that the IFR increased in the second wave.

